# Impact of Timing of and Adherence to Social Distancing Measures on COVID-19 Burden in the US: A Simulation Modeling Approach

**DOI:** 10.1101/2020.06.07.20124859

**Authors:** Oguzhan Alagoz, Ajay K. Sethi, Brian W. Patterson, Matthew Churpek, Nasia Safdar

## Abstract

**Background:** Across the U.S., various social distancing measures were implemented to control COVID-19 pandemic. However, there is uncertainty in the effectiveness of such measures for specific regions with varying population demographics and different levels of adherence to social distancing. The objective of this paper is to determine the impact of social distancing measures in unique regions.

**Methods:** We developed COVid-19 Agent-based simulation Model (COVAM), an agent-based simulation model (ABM) that represents the social network and interactions among the people in a region considering population demographics, limited testing availability, imported infections from outside of the region, asymptomatic disease transmission, and adherence to social distancing measures. We adopted COVAM to represent COVID-19-associated events in Dane County, Wisconsin, Milwaukee metropolitan area, and New York City (NYC). We used COVAM to evaluate the impact of three different aspects of social distancing: 1) Adherence to social distancing measures; 2) timing of implementing social distancing; and 3) timing of easing social distancing.

**Results:** We found that the timing of social distancing and adherence level had a major effect on COVID-19 occurrence. For example, in NYC, implementing social distancing measures on March 5, 2020 instead of March 12, 2020 would have reduced the total number of confirmed cases from 191,984 to 43,968 as of May 30, whereas a 1-week delay in implementing such measures could have increased the number of confirmed cases to 1,299,420. Easing social distancing measures on June 1, 2020 instead of June 15, 2020 in NYC would increase the total number of confirmed cases from 275,587 to 379,858 as of July 31.

**Conclusion:** The timing of implementing social distancing measures, adherence to the measures, and timing of their easing have major effects on the number of COVID-19 cases.

**Primary Funding Source:** National Institute of Allergy and Infectious Diseases Institute

## Introduction

The novel coronavirus disease 2019 (COVID-19) pandemic poses unprecedented challenges for communities and policy makers. SARS-CoV-2, the virus that causes COVID-19, is spread mainly through respiratory droplets and following contact with contaminated surfaces, but limited knowledge of transmission dynamics make effective control of the disease tailored to specific communities challenging. In the absence of an effective vaccine or treatment, non-pharmaceutical interventions, such as social distancing measures, which require one to maintain a 6-foot physical distance from non-household members, are the only means of reducing the spread of COVID-19. Examples include closures of schools, ceasing recreational activities, cancellation of large public gatherings, and shelter-in-place policies. When these measures are implemented and followed, daily counts of new COVID-19 cases decrease (“flattening the curve”).^1^ The negative economic and societal consequences of social distancing warrant a “dialing back” of such policies when it is safe to do so. However, the effect of easing social distancing on the transmission of SARS-CoV-2 is unclear. A set of indicators have been proposed by the current administration to guide communities on when they might consider easing rigorous mandated social distancing.^2^ However, these lagging indicators, such as hospitals having the capacity to “treat patients without crisis care”, are not ideal because once the number of new infections increases, re-implementation of social distancing is not as effective as a mitigation option due to exponential growth in the case of transmissible infectious diseases such as COVID-19.^2^ The ability to predict the effect of easing of social distancing is important to provide leading indicators for the right time to do this for a particular community.

Using the best information available, mathematical modeling of SARS-CoV-2 transmission dynamics allow scientists to forecast the effect of social distancing on the COVID-19 pandemic. In particular, agent-based models provide a flexible and simulation-based method to better represent transmission dynamics in a complex system. While other models are available, including those providing predictions for every state in the U.S., these models have several limitations that severely limit their generalizability.^3-7^ For example, they assume a closed population and ignore imported infections into the region, do not consider imperfect adherence levels to the dynamic social distancing measures accurately, and most do not incorporate the effect of limited testing capacity into the number of confirmed cases.^3-6^

We present an agent-based model that represents the social network and interactions among the people in a region considering local population demographics, population density, daily number of contacts in the absence of social distancing measures, and adherence to social distancing measures. Our model, referred to as COVid-19 Agent-based simulation Model (COVAM), allows transmission from asymptomatic patients, allows both community spread and transmission in the hospitals, accounts for the “imported” cases at the early days of the pandemic, and considers the possibility that some patients with mild to moderate symptoms never receive confirmatory testing for COVID-19. This study demonstrates how COVAM can inform decision-making in three unique urban communities as to how social distancing measures might be adjusted to control the spread of SARS-CoV-2 and prevent a return to exponential growth in the number of COVID-19 cases.

## Methods

We use an agent-based simulation modeling approach, which allows representing heterogeneous individuals who can behave independently. Individuals in COVAM have unique attributes, such as age and interact with each other through which SARS-CoV-2 is transmitted. We use a time step of one simulated day to update the status of the individuals and represent interactions. We assume all individuals in the model are susceptible to COVID-19 at the beginning of the simulation, i.e., no vaccination is available and there is no pre-existing immunity. We provide the details of the modeling approach and parameter estimation in Appendix A and only briefly summarize them here.

All individuals in COVAM are categorized into one of eight possible states representing an individual’s COVID-19-related status (Figure 1). We adopt our states using the clinical states as described by the Centers for Disease Control and Prevention (CDC) and a previous model.^3,8^ We consider transmission by patients in the exposed state during the last several days of the incubation period and allow some patients to never be tested positive for COVID-19 even when they experience mild symptoms, which reflects limited testing capacity in the earlier days of the pandemic in the U.S.^9-12^

**Figure 1.**
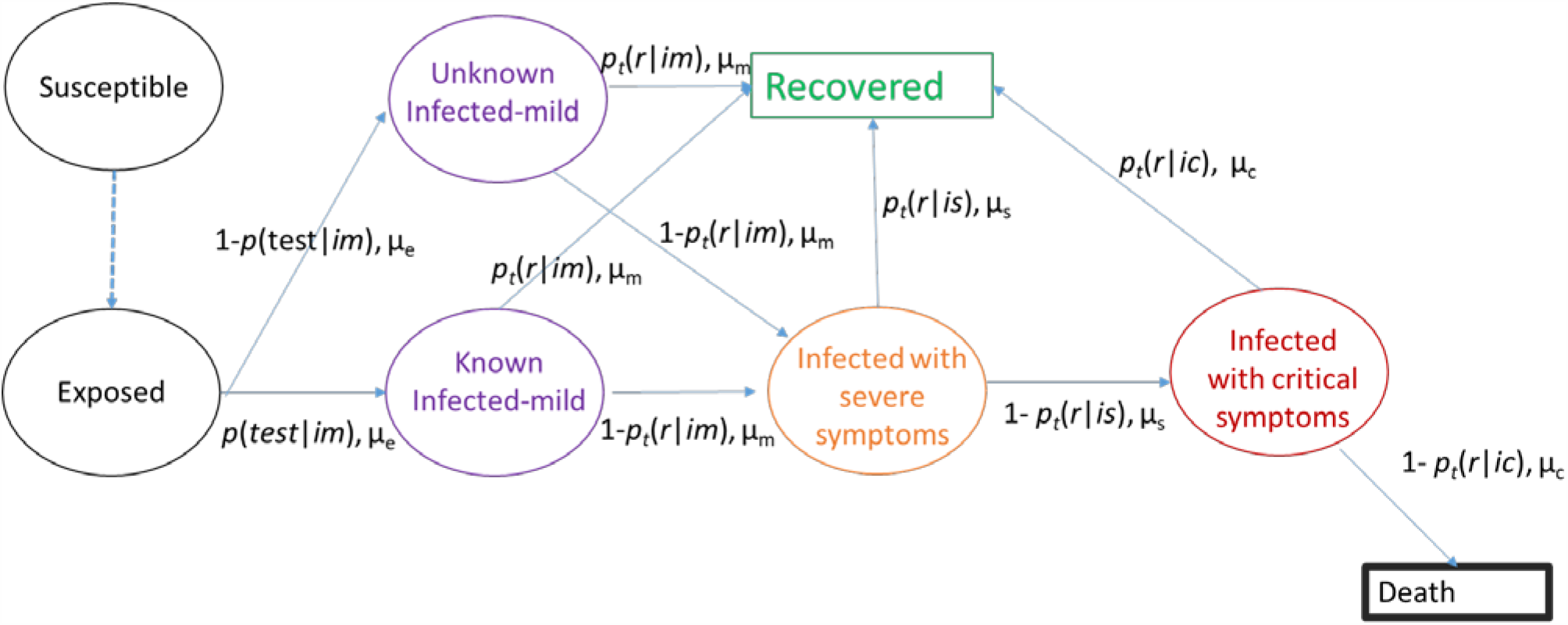
Progression of COVID-19 in the individuals.

The simulation starts with one (or more) exposed individual(s). At the beginning of each day, contagious individuals randomly interact with a number of other individuals in the community. For each interaction, there is a possibility that the contagious individuals expose the susceptible individual to SARS-CoV-2.

### Input Parameters

To maximize model generalizability, we derived input parameter estimates from relevant results in peer-reviewed literature and used data from Dane County, Wisconsin to calibrate several parameters (Table 1). COVAM has two different sets of parameters governing the transmissibility of SARS-CoV-2:

**Table 1.** List of the input parameters.

| Name | Description | Notation | Mean Value | Source |
| --- | --- | --- | --- | --- |
| **Parameters related to progression and duration** |  |  |  |  |
| Incubation period | Time between exposure until symptoms occur | $\mu_e$ | 5 days | CDC and literature<sup>8,29-31</sup> |
| Duration of mild symptoms | Time between the beginning of the mild symptoms and beginning of the severe symptoms (or recovery) | $\mu_m$ | 6 days | CDC and literature<sup>3,8,32,33</sup> |
| Duration of severe symptoms (hospitalization) | Time between the beginning of the severe symptoms and beginning of the critical symptoms (or recovery) | $\mu_s$ | 6 days | CDC and literature<sup>3,8,32,33</sup> |
| Duration of critical symptoms (ICU stay) | Time between the beginning of the severe symptoms and death or recovery | $\mu_c$ | 10 days | CDC and literature<sup>3,8,32-34</sup> |
| Probability of recovery after experiencing mild symptoms | Probability that an infected patient with mild symptoms (who also has tested positive) will recover once the mild symptomatic phase is over. This parameter is a function of age. | $p_t(r/im)$ | Age<br>$p_t(r/im)$<br>0–19 98%<br>20–44 82%<br>45–54 75%<br>55–64 75%<br>65–74 64%<br>75–84 55%<br>≥85 49% | CDC<sup>35</sup> |
| Probability of recovery after experiencing severe symptoms | Probability that an infected patient with severe symptoms will recover once the severe symptomatic phase is over. This parameter is a function of age. | $p_t(r/is)$ | Age $p_t(r/is)$<br>0–1 >99%<br>20–44 82%<br>45–54 68%<br>55–64 69%<br>65–74 63%<br>75–84 53%<br>≥85 65% | CDC<sup>35</sup> |
| Probability of recovery after experiencing critical symptoms | Probability that an infected patient with critical symptoms will recover once the critical symptomatic phase is over. This parameter is a function of age. | $p_t(r/is)$ | Age $p_t(r/is)$<br>0–19 >99%<br>20–44 95%<br>45–54 92%<br>55–64 75%<br>65–74 72%<br>75–84 64%<br>≥85 9% | CDC and Wisconsin Department of Health Services<sup>35,36</sup> |
| Probability that an infected patient with critical symptoms needs ventilator support | Probability that an infected patient with critical symptoms will need mechanical ventilator support during their ICU stay | $p(v/is)$ | 46% | Literature<sup>29</sup> |
| **Transmission parameters** |  |  |  |  |
| Number of contacts per person without any social distancing intervention | Number of people that an individual has a contact that likely produces a transmission opportunity. This parameter represents the number of such contacts. This parameter assumes no intervention is implemented. | $n_c$ | 10 | Calibration and literature<sup>37,38</sup> |
| Number of contagious days of an exposed patient | Exposed individuals could be contagious prior to showing any symptoms of the disease. This parameter represents the number of days in the end of incubation period and prior to showing mild symptoms when exposed patients transmit the disease. | $d$ | 2 | Literature<sup>12,39</sup> |
| Probability that an exposed patient will transmit SARS-CoV-2 to a susceptible individual with a close contact | Probability that an asymptomatic patient within the last few days of the incubation period will successfully transmit SARS-CoV-2 to a susceptible patient who is in close contact | $p(c\vert e)$ | 0.0418 | Calibration and literature<sup>13-15</sup> |
| Probability that a patient with mild to moderate symptoms and not tested positive will transmit SARS-CoV-2 to a susceptible individual with a close contact | Probability that a patient with mild to moderate symptoms and not tested positive will successfully transmit SARS-CoV-2 to a susceptible patient who is in close contact. | $p(c\vert im-)$ | 0.0418 | Calibration and literature<sup>13-15</sup> |
| Probability that a patient with mild to moderate symptoms and tested positive will transmit SARS-CoV-2 to a susceptible individual with a close contact | Probability that a patient with mild to moderate symptoms and tested positive will successfully transmit SARS-CoV-2 to a susceptible patient who is in close contact. | $p(c\vert im+)$ | 0.0418 | Calibration and literature<sup>13-15</sup> |
| Relative transmissibility of a patient with severe symptoms compared to a patient with mild to moderate symptoms and tested positive | This parameter describes the probability that a patient with severe infections transmits the disease ($p(c\vert is)$) relative to that for a patient with mild to moderate symptoms and tested positive transmitting the disease $p(c\vert im+)$. | $\frac{\ln(1 - p(c\vert is))}{\ln(1 - p(c\vert im+))}$ | 0% | Not applicable |
| Relative transmissibility of a patient with critical symptoms compared to a patient with mild to moderate symptoms and tested positive | This parameter describes the probability that a patient with critical infections transmits the disease ($p(c\vert ic)$) relative to that for a patient with mild to moderate symptoms and tested positive transmitting the disease $p(c\vert im+)$. | $\frac{\ln(1 - p(c\vert ic))}{\ln(1 - p(c\vert im+))}$ | 0% | Not applicable |
| **Diagnostic testing** |  |  |  |  |
| Baseline probability of testing with mild to moderate symptoms | The baseline probability that a patient who experiences mild to moderate symptoms will be tested positive with COVID-19, representing limited testing capacity and cases where some patients do not feel mild symptoms to make them request for testing, additional testing capacity increases this probability | $p(test\vert im)$ | 75% | Literature and calibration<sup>40,41</sup> |

1. Number of close contacts per day without any intervention, which represents the social network effect and is independent of the respiratory agent that is transmitted. This parameter depends on population density and is estimated using literature and calibration as explained below.
2. Probability that contagious exposed patients transmit SARS-CoV-2 to a susceptible individual when a close contact occurs.

The theoretical basic reproduction number (R0) corresponding to these parameter estimates is 3.34 for Dane County without any social distancing measures, which was within the range of R0 values reported in the literature (1.5 to 6.5).^13,14^ In particular, a recent study based on the COVID-19 epidemic in Italy reported an R0 value of 3.47 for the early days of the epidemic.^15^ Similarly, a recent study estimated the median R0 value for the Wuhan region as 5.7.^16^ Therefore, we concluded that our transmission parameters were within acceptable ranges.

### Adherence to Social Distancing Measures

The effectiveness of social distancing measures depends on how closely a population follows such measures and the type of measures that are implemented at different times. For example, in the state of New York, mass gathering restrictions started on March 12, 2020, initial business closures were recommended on March 16, 2020, educational facilities were closed on March 18, 2020, and, finally, non-essential services closed along with a statewide stay-at-home order on March 22, 2020.^5^ COVAM represents the adherence to social distancing explicitly by adjusting the number of contacts per person using cell-phone mobility data published by several sources.^17-19^ For instance, the average number of daily close contacts per person in New York City (NYC) is estimated as 20, therefore, a 70% adherence level reduces the number of such contacts to 6 per day per person leading to the slowing of transmission. Note that adherence to social distancing measures in COVAM is a proxy for several behaviors that reduce the transmissibility of SARS-CoV-2, including less frequent traveling, keeping at least 6-feet distancing during person-to-person interactions as well as frequent hand washing and wearing masks.

### Calibration and Validation

Several model input parameters involve a high level of certainty, including disease transmission rates, probability of testing for COVID-19, and adherence to social distancing measures. We used a simple calibration procedure using earlier surveillance data from Dane County to fine-tune these parameters (Table 1). We used the reported data for COVID-19 from Dane County until May 15, 2020 to test whether our initial parameter estimates replicated the number of cases accurately. We did not change any of the parameters except the adherence to social distancing input after May 15, 2020. We stopped adjusting adherence input as of May 15, 2020 for NYC and May 26, 2020 for Dane County and Milwaukee compared the model’s projections to actual number of cases after this date.

### Application to Dane County

Input parameters that are used for the computational experiments for Dane County are presented in Table 2. Briefly, we incorporated the population demographics in terms of age groups, number of individuals who are “imported” into Dane County from outside of Dane County, and adherence to social distancing measures. The model has the ability to add different numbers of imported cases on a daily basis, however we kept the number of initial imported cases the same to prevent overfitting. We considered that adherence to social distancing measures in Dane County dropped on May 14, 2020 since Wisconsin Supreme Court struck down the stay-at-home order of the governor in the state of Wisconsin on this date.^20^ We assumed that adherence to social distancing remains the same after May 21, 2020.

**Table 2.** Input parameters used to apply COVAM to Dane County, Milwaukee, and NYC.

| Name | Description | Value for Dane County | Value for Milwaukee* | Value for NYC | Source |
| --- | --- | --- | --- | --- | --- |
| Population | Number of people living in the region | 542,364 | 1,576,236 | 8,398,748 | US census data<sup>42</sup> |
| Simulation start date | The date when the simulation starts | March 4, 2020 | March 4, 2020 | March 4, 2020 | Not applicable |
| Number of initial exposures | The number of individuals who were exposed to COVID-19 at the beginning of the simulation | 1 | 3 | 16 | Calibration |
| Number of imported cases and dates | Number of individuals who are exposed to COVID-19 from outside of the region per day and the dates for importing cases | 3 per day between March 5, 2020 and March 25, 2020, 1 per day afterwards | 6 per day between March 5, 2020 and April 3, 2020, 3 per day afterwards | 160 per day between March 5, 2020 and April 3, 2020, 80 per day afterwards | Calibration |
| Number of close contacts per person per day without any social distancing intervention | Number of people that an individual has a contact that produces a transmission opportunity. This parameter assumes that the schools are closed. | 10 | 10 | 20 | Calibration and literature<sup>37,38</sup> |
| School effect | % increase in the number of daily close contacts when the schools are open | 40% | 40% | 40% | Literature<sup>37</sup> |
| Adherence to social distancing | % of individuals who are following the social distancing guidelines, which is equivalent to a % drop in average number of contacts per person | Mar 4-11: 0%<br>Mar 12-25: increasing linearly from 0% to 90%<br>Mar 26-April 30: 80%<br>May 1-13: 70%<br>May 14-20: 60%<br>May 21-end of simulation: 70% | Mar 4-11: 0%<br>Mar 12-22: increasing linearly from 0% to 60%<br>Mar 23-Apr 7: 60%<br>Apr 8-12: 80%<br>Apr 13-May 13: 70%<br>May 14-25: 60%<br>May 26-end of simulation: 70% | Mar 4-11: 0%<br>Mar 12-20: increasing linearly from 0% to 80%<br>Mar 21- Apr 12: 80%<br>Apr 13- end of simulation: 90% | Mobile phone data and calibration<sup>17-19</sup> |
| Probability of testing | The probability that a patient who experiences mild to moderate symptoms will be tested positive with COVID-19 | 75% until April 15, 80% between April 15 and May 10, 90% afterwards | 75% until April 15, 80% between April 15 and May 10, 90% afterwards | 75% until April 15, 80% between April 15 and May 10, 90% afterwards | Literature and calibration<sup>40,41</sup> |
| Demographics | % of the individuals in different age groups | Age Prop.<br>0–19 20.8%<br>20–44 46.5%<br>45–54 10.3%<br>55–64 10.2%<br>65–74 7.4%<br>75–84 3.3%<br>≥85 1.5% | Age Prop.<br>0–19 28.6%<br>20–44 36.2%<br>45–54 13.7%<br>55–64 8.5%<br>65–74 6.6%<br>75–84 4.7%<br>≥85 1.0% | Age Prop.<br>0–19 23.2%<br>20–44 38.6%<br>45–54 13.0%<br>55–64 11.7%<br>65–74 7.1%<br>75–84 3.4%<br>≥85 1.9% | US census data<sup>42</sup> |

\*Milwaukee Metro area consists of four counties, including Milwaukee County, Waukesha County, Washington County, and Ozaukee County; Prop.= proportion of the population in this age group.

**Table 3.** Impact of easing social distancing measures on the total number of confirmed cases in different dates in NYC.

| Total number of infections by | Easing social distancing measures on June 1 |  | Easing social distancing measures on June 15 |  | Easing social distancing measures on July 1 |  |
| --- | --- | --- | --- | --- | --- | --- |
| NYC |  |  |  |  |  |  |
|  | DAE 10% | DAE 20% | DAE 10% | DAE 20% | DAE 10% | DAE 20% |
| June 30 | 239,605 | 325,510 | 218,096 | 224,575 | 213,910 | 213,910 |
| July 31 | 379,858 | 2521140 | 275,587 | 711,792 | 235,866 | 287,118 |
| August 31 | 807,300 | 5,735,160 | 480,114 | 4,576,610 | 328,531 | 1,834,100 |

DAE: Drop in adherence rates after easing social distancing measures. A value of 10% and 20% DAE implies that adherence to social distancing measures after the date of easing is at 70% and 60%, in NYC, respectively.

### Application to Milwaukee Metropolitan Area

We adapted COVAM to Milwaukee to cross-validate our model and test its predictive accuracy. Our objective was to modify as few parameters as possible to prevent overfitting. We used the same simulation settings as those in Dane County except four changes (Table 2): 1) epidemic was initiated with three exposed patients instead of one exposed patient; 2) six imported cases were added to account for the larger population; 3) population demographics using Milwaukee population was adjusted; and 4) adherence to social distancing measures was adjusted proportionate to cell phone mobility data which indicated lower adherence in Milwaukee as compared to Dane County.^17-19^ We assumed that adherence to social distancing remains the same after May 26, 2020.

### Application to New York City

New York City, NY (NYC) was among the first epicenters for the COVID-19 pandemic in the U.S. Thus, the relative maturity of the epidemic in NYC made it a good test case to evaluate COVAM’s predictive accuracy for later stages of the pandemic. As before, to maximize generalizability and avoid overfitting, we used the same simulation settings as those in Dane County except four changes (Table 2): 1) 160 imported cases were added between March 5, 2020 and April 4, 2020 and 80 imported cases were added after April 4, 2020 to account for the greater number of visitors to NYC; 2) population demographics using the NYC population information was adjusted; 3) adherence to social distancing measures was adjusted, as mobile phone data showed that adherence was higher in NYC as compared to Dane County;^17-19,21^ and 4) number of contacts per person in each day was set to 20 due to significantly higher population density (438/square mile in Dane County vs. 27,755/square mile in NYC), as social network analysis literature reports that population density increases the number of close contacts.^22,23^ These parameters correspond to a theoretical R0 value of 6.68.

### Policy Analyses

We used COVAM to evaluate the impact of three different aspects of social distancing:

1. *Adherence to social distancing measures.* We compared the case that social distancing is not implemented (schools are open/closed) to the case that social distancing is implemented at the beginning of the simulation, and adherence to social distancing is consistently at a level of 25%-50%-75%-90%.
2. *Timing of implementing social distancing.* We tested the scenario that social distancing is implemented 1 week earlier and 1-4 weeks later than the actual implementation date. We assumed that social distancing measures in NYC end on June 1 and adherence thereafter does not become zero but drops 20 percentage points due to heightened awareness of the community. In NYC, a drop of 20 percentage points in adherence to social distancing reduces the adherence level from 90% to 70% after easing social distancing measures. For Dane County and Milwaukee, we did not change adherence to social distancing after May 21, 2020 and May 26, 2020, respectively.
3. *Timing of easing social distancing.* We tested the scenario that social distancing measures are eased on June 1, June 15, and July 1. We assumed that after social distancing measures are eased, due to the heightened awareness of the community, adherence to social distancing drops by 10 and 20 percentage points compared to the adherence levels before easing social distancing measures in each region. For example, in NYC, a drop of 10 percentage points in adherence to social distancing reduces the adherence level from 90% to 80% after social distancing measures are eased. This is consistent with the adherence levels observed in Dane County and Milwaukee after Wisconsin Supreme Court struck down the stay-at-home order of the governor in the state on May 13, 2020.^20^ We conducted these experiments only for NYC.

We conducted a sensitivity analysis for NYC region on two parameters: probability of testing given a patient experiences mild to moderate symptoms and transmission rates. Testing has been severely limited especially in the early days of epidemic in the U.S., therefore, it is likely that our initial calibration may not have estimated the input parameters correctly. For example, recent data from NYC suggest that one of every five residents tested positive for antibodies for COVID-19.^24^ For this purpose, we replaced our base estimate of 75% for the probability of testing with 25% and 50% and recalibrated model parameters and reevaluated all of the scenarios described above. Similarly, we reduced the transmission rate by 50% (from a base estimate of 0.0418 to 0.0209 for the probability of transmission from a patient experiencing mild to moderate symptoms to a susceptible patient when a close contact occurs), recalibrated model parameters, and reevaluated all of the scenarios. We ran 100 replications for each experiment and report only mean values since the standard errors were very low.

## Results

COVAM replicated the observed number of COVID-19 cases, and thus SARS-CoV-2 transmission dynamics, over time in the short term accurately (Figure 2). Our first set of experiments showed that adherence to social distancing has a significant effect on the cumulative number of cases (Figure 3 and Appendix Table 1). For example, compared to social distancing at a 50% adherence level, no social distancing while closing schools increases the total number of cases from 55,592 to 480,630 in NYC in just 26 days (by March 31).

**Figure 2.**
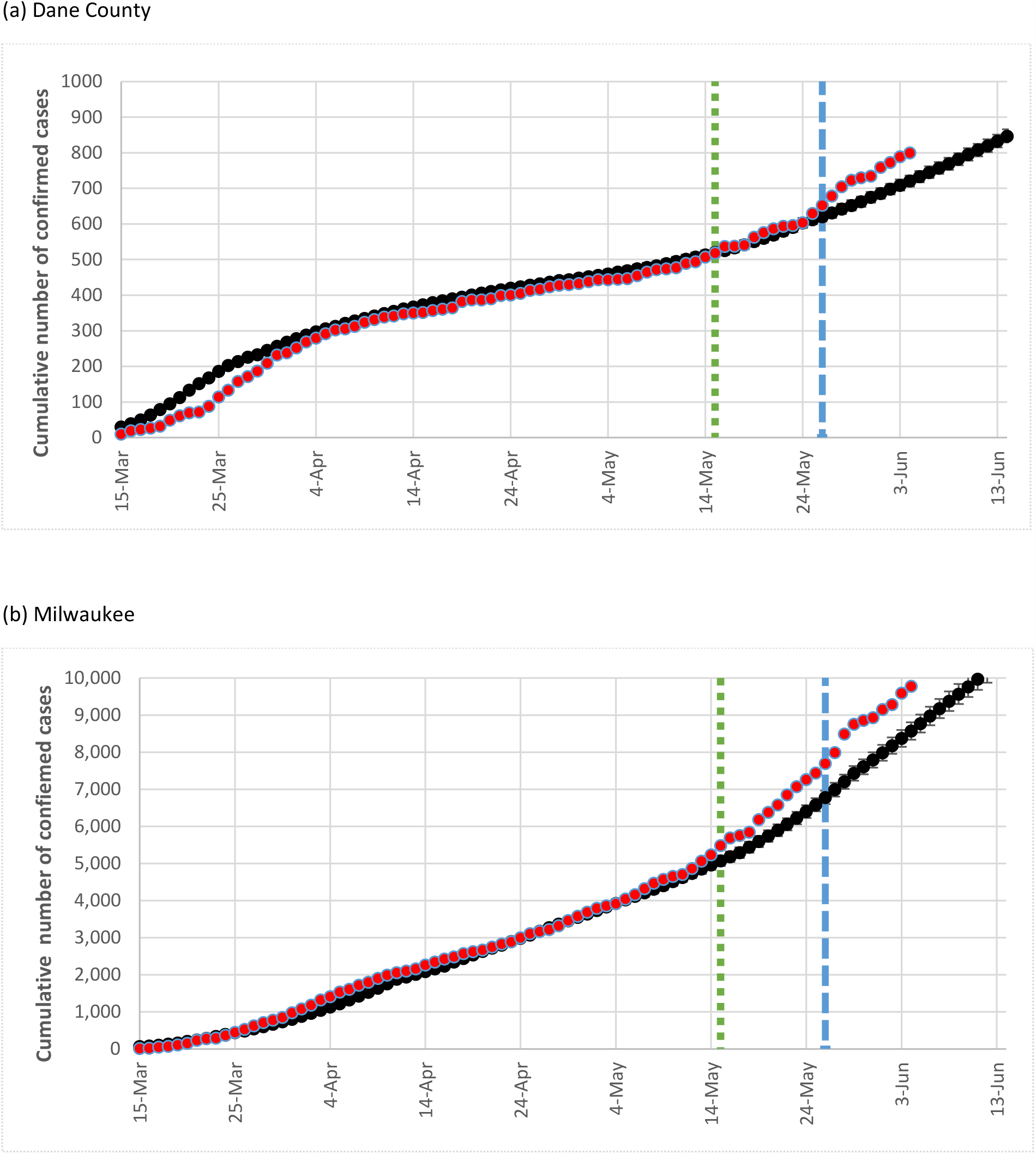
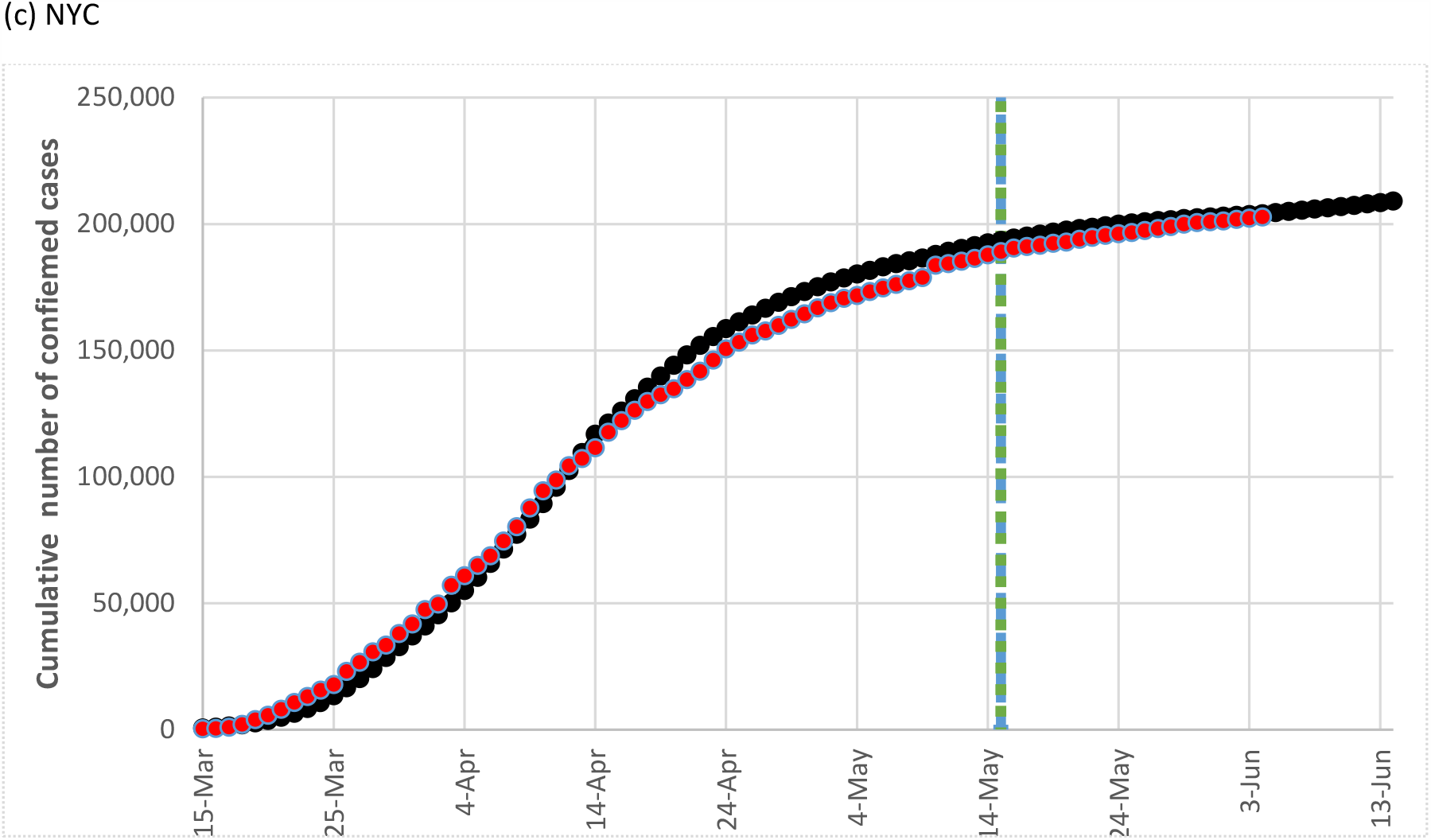
Model validation results for the base case. In each of the following figures, red dots represent actual observed cumulative number of confirmed cases, black solid line represents the model’s predictions, and error bars around the black solid line represent 95% confidence intervals for the model’s predictions based on 100 replications, green dotted line represents the date after which all model input parameters were fixed except adherence to social distancing measures, and blue dashed line represents the date after which no model input parameter was modified.

**Figure 3.**
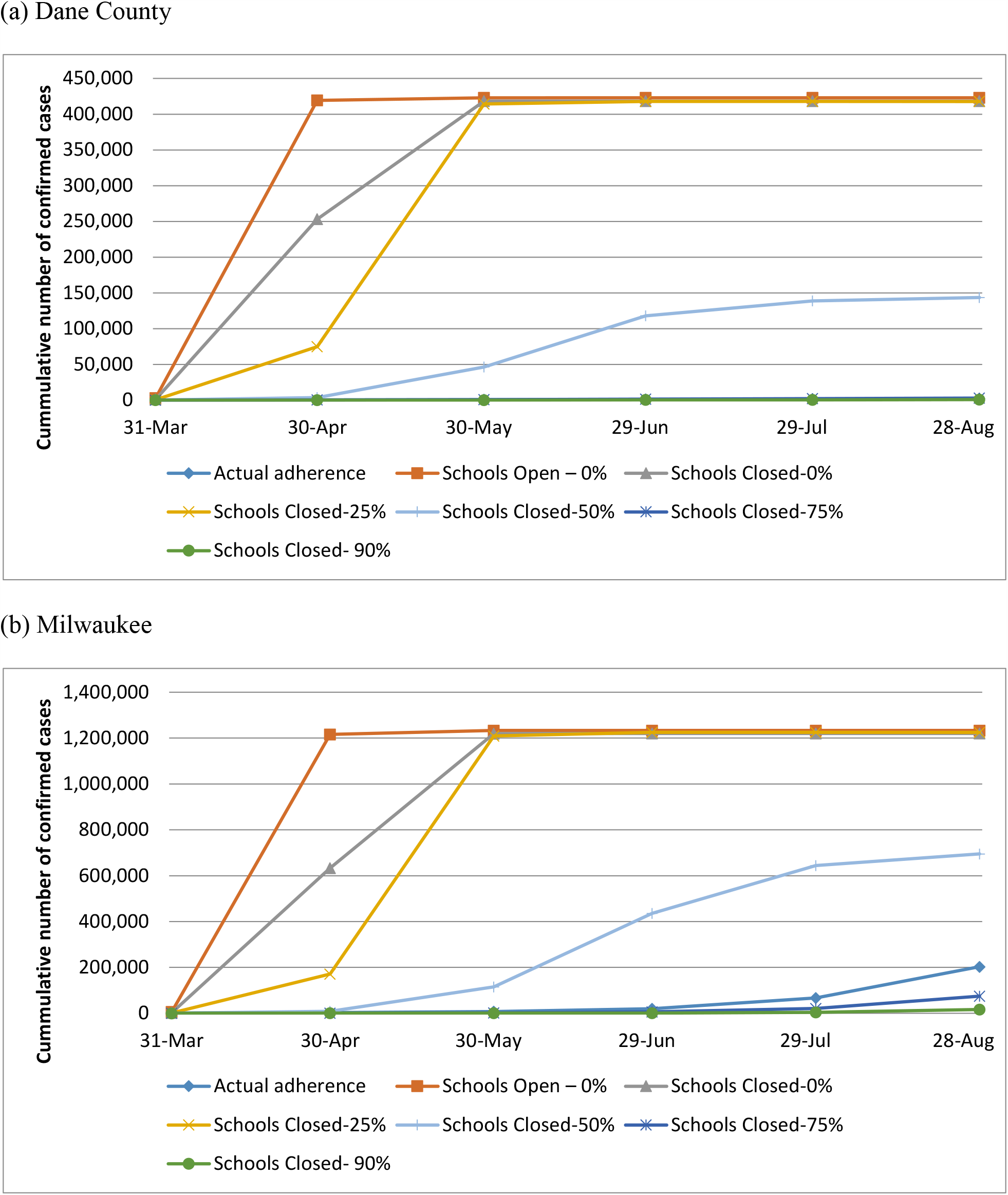
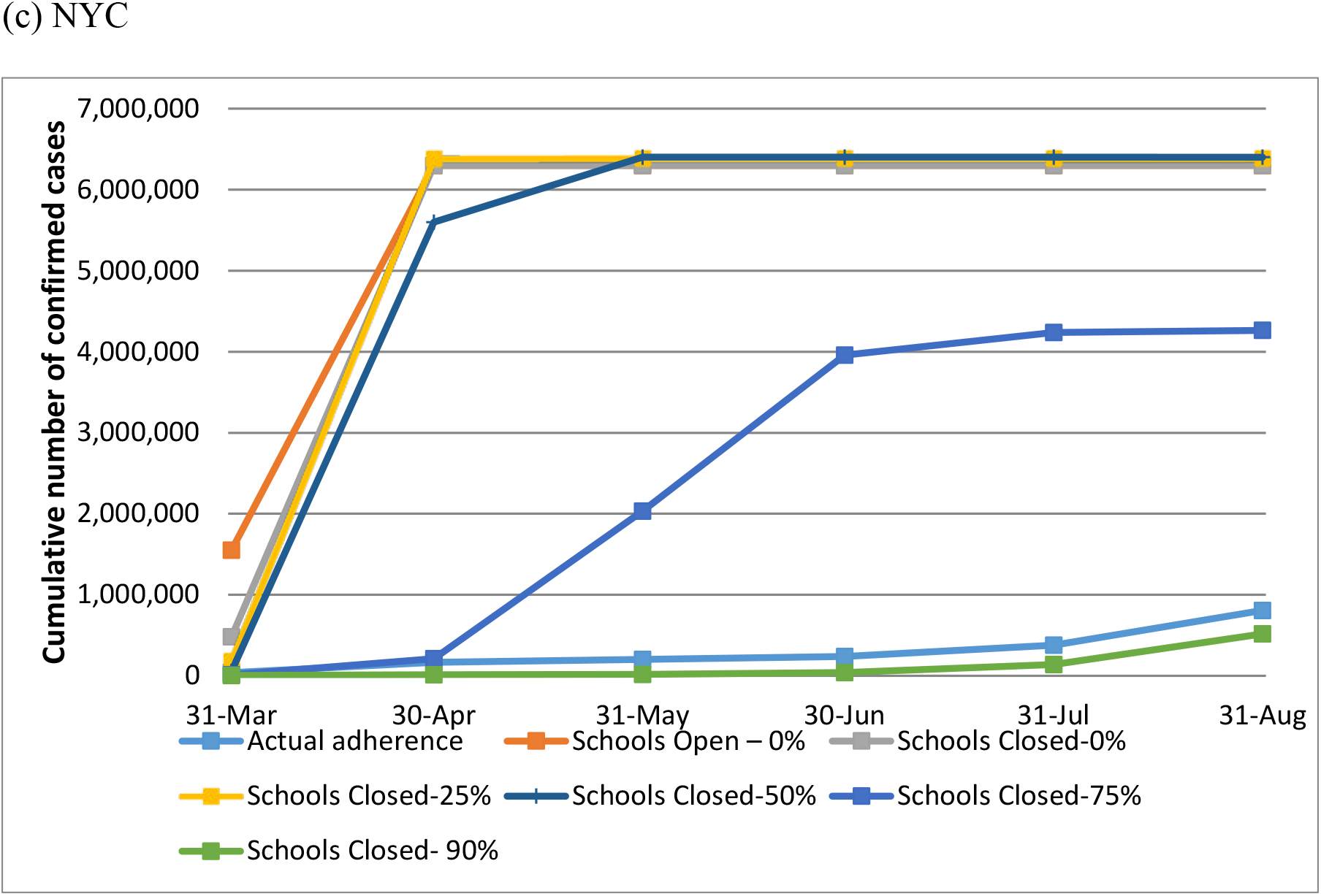
**Impact of adherence to social distancing on the total number of confirmed cases in different dates (a) Dane County (b) Milwaukee (c) NYC**

Figure 4 presents how various outcomes associated with COVID-19 would change when social distancing measures are implemented earlier or later than the actual implementation date. We found that even a 1-week delay in implementing social distancing interventions would have had a significant impact on the total number of confirmed infections over time in each of the three urban communities (Figure 4). For example, implementing social distancing measures one week earlier in NYC would have reduced the number of cases by 77% from 191,984 to 43,968 by May 30 whereas a 1-week delay in initiating such measures could have increased the number of confirmed cases by more than six-fold to 1,299,420 (Figure 4 and Appendix Table 2). The impact of implementing social distancing measures depends highly on the region as each region has different levels of adherence to social distancing and are experiencing different levels of transmission (Appendix Table 2). For example, implementing the measures on March 19 instead of March 12 in Dane County increases the number of cases by 240% as of May 15 (498 to 1693), while the same scenario in NYC increases the number of cases by 576% (191,984 to 1,299,420). These results demonstrate the differential impact of implementing social distancing measures in urban communities. A delay in implementation has a differential effect on the number of cases in the Milwaukee as compared to Dane County, indicating that the impact of a delay in implementing social distancing measures varies significantly even within the same state.

**Figure 4.**
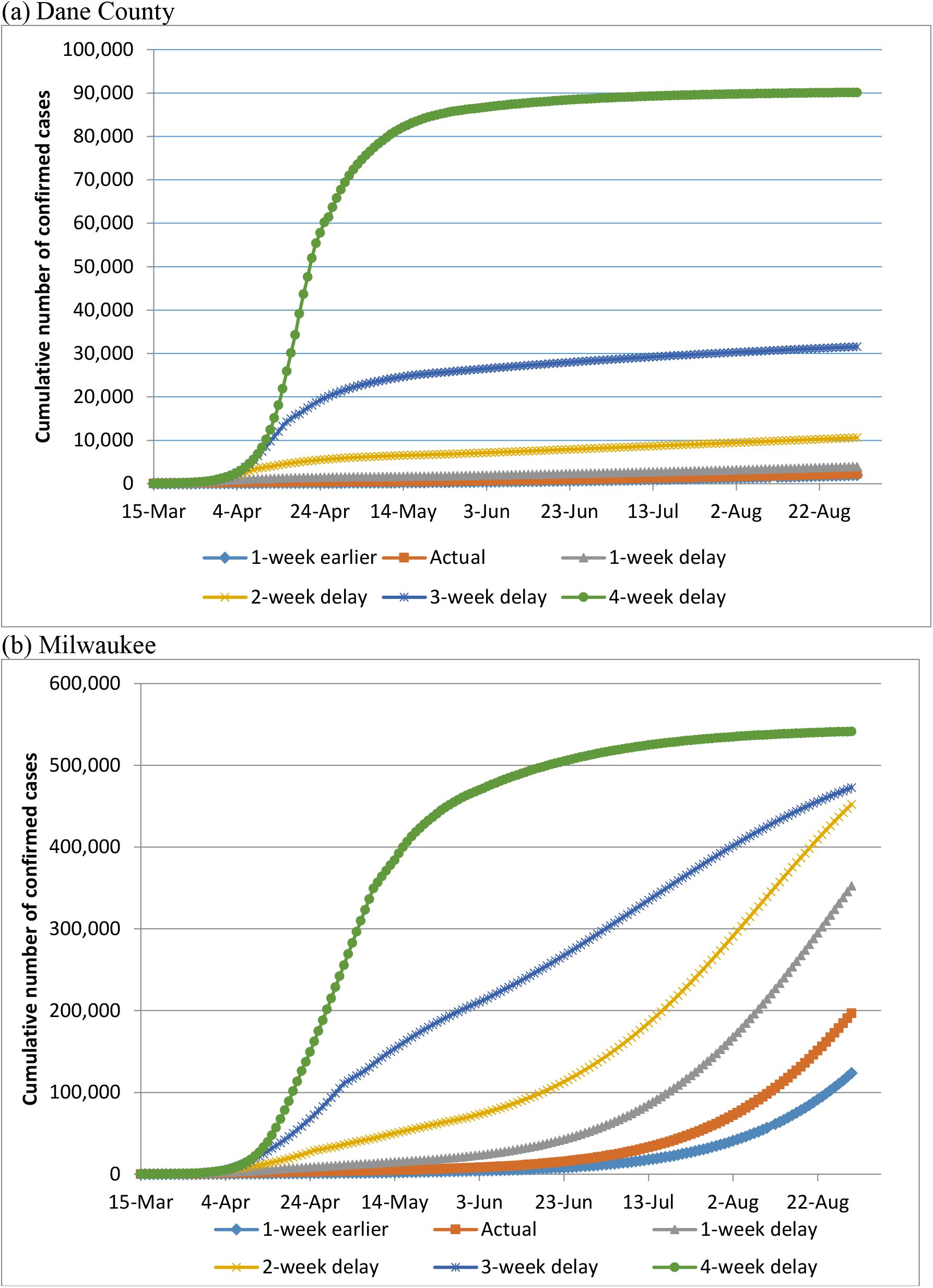
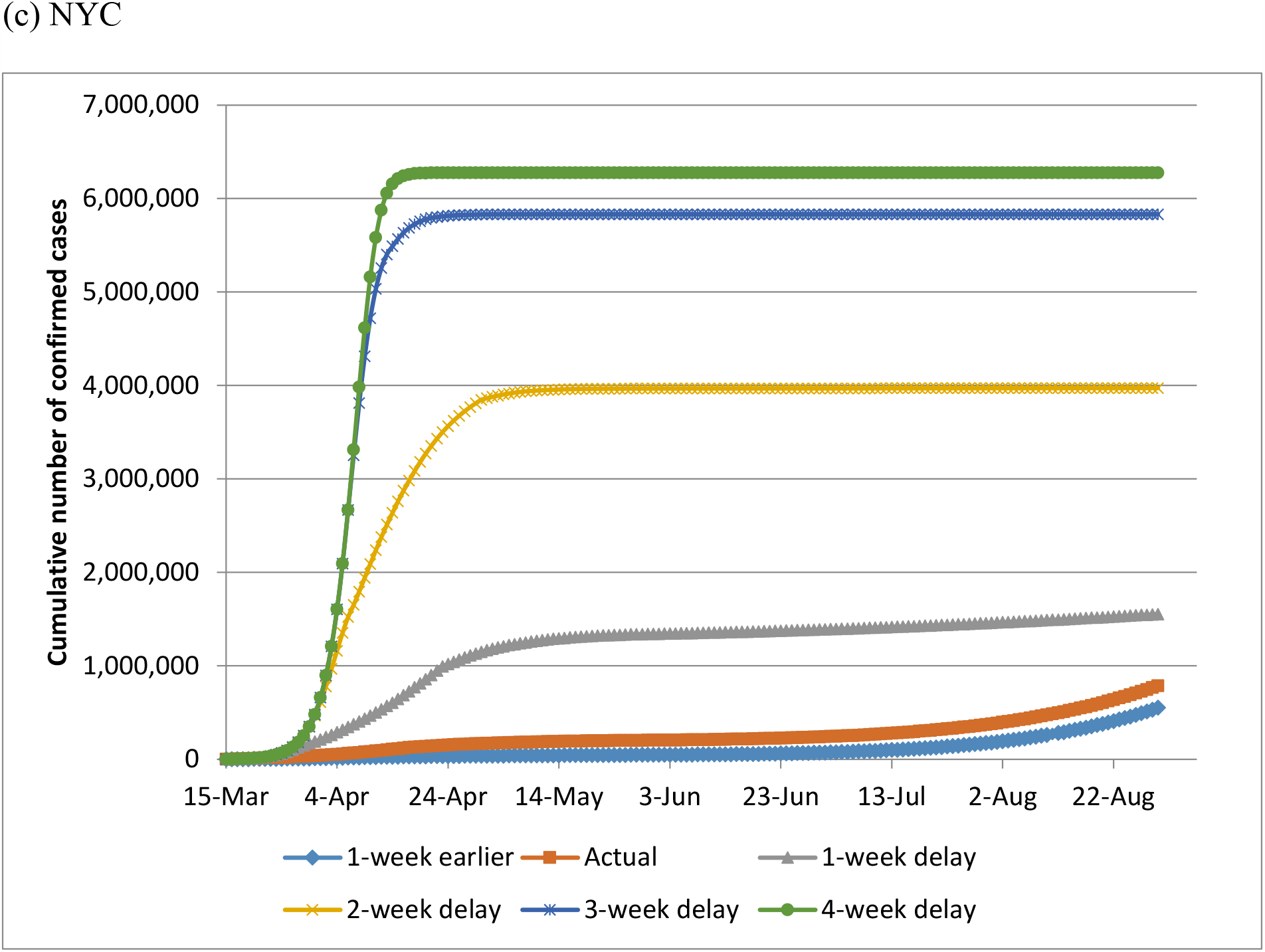
**Comparison of total number of confirmed cases over time for implementing social distancing in different dates (a) Dane County (b) Milwaukee (c) NYC**

Table 2 shows that earlier easing of the social distancing measures would have major detrimental effects on the total numbers of COVID-19 cases in NYC. Premature easing of social distancing measures on June 1 instead of June 15 would increase the total number of confirmed cases from 275,587 to 379,858 as of July 31. Table 2 also shows that in NYC, after easing social distancing measures on June 15, increasing the adherence level by just 10 percentage points would reduce the number of cases from 711,792 to 275,587 as of July 31, demonstrating the importance of practicing personal behaviors that prevent the transmission of SARS-CoV-2 after the social distancing measures are eased.

The sensitivity analysis on the probability of testing and transmission rates showed that even if a lower probability of testing or a lower rate of transmission were to be used, the overall trends in base-case runs would still hold (Appendix Figures 1-6, and Appendix Tables 3-11).

## Discussion

In this simulation study, we estimated the impact of the timing of implementing and easing of and adherence to social distancing measures in three unique urban communities using agent-based simulation modeling. We found that the timing of implementing social distancing and adherence to social distancing significantly affected the number of cases. The impact of the timing of implementation of social distancing measures varied widely by region. In NYC, the impact was large compared to Dane County and Milwaukee. This finding illustrates the importance of considering implementing and reopening policies at the regional level, as the results in Dane County and Milwaukee differed considerably despite being in the same state. We further found keeping a high level of adherence after easing social distancing measures has a major impact on the number of cases, implying that cities and regions should strongly encourage the community to maintain behaviors that reduce the transmissibility of SARS-CoV-2 such as wearing masks. The accuracy of COVAM for predicting the current outbreak and its ability to estimate the impact of easing social distancing measures at the regional level demonstrates its unique value for informing current and future policies. Our findings clearly show that one-size-fits-all strategies are suboptimal and that context- and region-specific policies are needed when considering implementing and easing social distancing measures.

Some opponents of stay-at-home policies are motivated by the prospects of achieving herd immunity as a reason for easing current social distancing measures that are in place.^25^ However, herd immunity is only possible if infection with SARS-CoV-2 results in solid immunity, which is unknown at this time. Moreover, the correlates of protective immunity to SARS-CoV-2 have yet to be identified.^25^ In the absence of this understanding, social distancing is the most effective tool to prevent further spread of SARS-CoV-2. To date, social distancing measures and adherence to those measures have resulted in halting exponential growth in the daily case counts of COVID-19. However, SARS-CoV-2 continues to spread and major cities and urban communities in the U.S. have yet to return to pre-exponential growth levels of transmission. COVAM demonstrates that premature easing of social distancing measures and adherence to these measures will result in rapid return to exponential growth of COVID-19 cases and negate progress made to date in slowing the spread of SARS-CoV-2 in communities, managing the healthcare capacities of health systems, and preventing larger numbers of deaths.

Our study has limitations, most of which are due to limited available data and uncertainty regarding SARS-CoV-2. We make simplifying assumptions such as asymptomatic patients transmit the disease at the same rate as symptomatic patients and weather does not affect SARS-CoV-2 transmissibility whereas several studies suggest otherwise.^26,27^ Furthermore, COVAM uses adherence to social distancing as a proxy for several factors contributing to the disease transmission including fewer close contacts due to limited travel and precautions that prevent transmission during a close contact such as wearing masks, therefore COVAM may not be accurately estimating the impact of personal precautions on transmission. To overcome this limitation, we assumed a high level of adherence after the easing of social distancing measures to provide a conservative estimate on the effect of easing social distancing measures.

There are also several limitations related to the modeling approach. Our calibration procedure used a simple trial-error approach as opposed to a full-scale calibration in which all possible combinations of the input parameter values within a plausible range are tested.^28^ Due to the computational intensity of a more formal and detailed calibration procedure, our calibration process may have not identified the best parameter combinations to represent the pandemic.

Additionally, we used mean parameter estimates instead of probability distributions for input parameters to reduce computational time, although we do not expect this simplification to substantively change our results.

In conclusion, our model demonstrates that the delayed implementation, lower adherence to, and premature easing of social distancing generally result in increased cases of COVID-19 in urban areas of the U.S. The magnitude of impact, however, varies significantly by region. These findings highlight the importance of region-specific considerations, and ideally modeling, as inputs to making policy decisions for a given region.

## Data Availability

All underlying data for this paper is included in the paper and the appendix.

## Acknowledgements

The authors are grateful for the early feedback received for development and model use during Covid-19 epidemic from researchers, policy makers, and stakeholders. In particular, the authors thank UW Health Data Science & Advanced Analytics group including Dr. Frank Liao, Croix Christenson, Corey Fritsch, Joel Galang, and Sabrina Adelaine. The authors also thank Amanda Young for her help in preparing this manuscript.

## Notes

### Competing Interest Statement

The authors have declared no competing interest.

### Funding Statement

This work was supported by the National Institutes of Health under National Institute of Allergy and Infectious Diseases Institute (NIAID) Grant 1DP2AI144244-01. The content is solely the responsibility of the authors and does not necessarily represent the official views of the National Institutes of Health.

### Author Declarations

This study is exempt for IRB.

